# Identification of SYT10 as a potential prognostic biomarker in esophageal cancer by comprehensive analysis of a mRNA-pseudogene/lncRNA-miRNA ceRNA network

**DOI:** 10.1101/2023.05.17.23290118

**Authors:** Milad Daneshmand-Parsa, Sharareh Mahmoudian-Hamedani, Parvaneh Nikpour

**Affiliations:** Department of Genetics and Molecular Biology, Faculty of Medicine, Isfahan University of Medical Sciences, Isfahan, Iran

**Keywords:** Biomarkers, Competitive endogenous RNA, Esophageal cancer, Pseudogene

## Abstract

**Background:** Esophageal carcinoma (ESCA) is often diagnosed at the advanced stages, has a poor survival rate and overall is one of the deadliest cancers world-wide. Recent studies have elaborated the significance of non-coding RNAs like pseudogenes, long non-coding RNAs (lncRNAs) and microRNAs (miRNAs) in cancer progression. In this study, we constructed a four-component competing endogenous RNA (ceRNA) network in ECSA and suggested an RNA with prognostic potential.

**Materials and methods:** Expression profiles of mRNAs, pseudogenes, lncRNAs and miRNAs were collected from The Cancer Genome Atlas (TCGA) database. A ceRNA network was constructed based on differentially-expressed RNAs. KEGG and GO functional analysis and PPI network analysis was carried out on differentially-expressed (DE) RNAs of the ceRNA network. Survival analysis was carried out on a selection of RNAs with the highest degree centrality ranks to discover potential prognostic biomarkers.

**Results:** A four-component ceRNA network with 529 nodes and 729 edges was constructed. The most significant GO biological process terms included signal transduction, cell adhesion and positive regulation of gene expression. The analysis of KEGG pathways showed that DEmRNAs were significantly enriched in pathways such as cytokine-cytokine receptor interaction and Cell cycle. Amongst the RNAs that were found to be associated with survival, *SYT10* had the highest hazard ratio and thus, proved to be a potential prognostic biomarker for ESCA.

**Conclusion:** Our study presented a four-component ceRNA network for ESCA, and identified RNA candidates that were associated with survival of ECSA. Further experimental evaluations and precise validation studies are needed for their clinical significances and roles in the progression of ESCA.

## Introduction

Esophageal cancer (ESCA) is the eighth common cancer world-wide with Asia leading with highest rates of incidence and mortality (Malhotra *et al*., 2017), and shows a sharp increase in incidence in other parts of the globe, too (Lepage *et al*., 2008). The disease inflicts men more than women (Malhotra *et al*., 2017) and the outcome is also associated with sex, as females show significantly better outcome than males (Bohanes *et al*., 2012). ESCA is often diagnosed in the advanced stages (Chela *et al*., 2022), and because of its aggressiveness and poor survival rate, it is considered as one of the deadliest cancers in the world (Lepage *et al*., 2008). The overall 5-year-survival of ESCA patients is about 15-25%, and better prognosis is expected if the disease is detected at early stages (Pennathur *et al*., 2013).

Non-coding RNAs (ncRNAs) have proved to have impact on molecular mechanisms of ESCA tumorigenesis. One of the trends of the recent years’ studies is to investigate the potential impact of ncRNAs in tumorigenesis through analyzing competing endogenous RNA (ceRNA) networks (Guo *et al*., 2021; Donyavi, Salehi-Mazandarani and Nikpour, 2022; Qiu *et al*., 2022). Coding and non-coding RNAs that share the same miRNA response elements (MREs) are able to compete with each other to bind to the same miRNAs and referred to as ceRNAs (Salmena *et al*., 2011). As a result of this competition, a network of ceRNAs is created which dysregulation of components is associated with tumor formation and invasion (Ala, 2021; Yang *et al*., 2021).

One of the main components of ceRNA networks are pseudogenes. Pseudogenes are defective copies of genes spread across the genome (Cheetham, Faulkner and Dinger, 2020) which used to be regarded as genomic fossils with no apparent functions (Chen *et al*., 2020). However, recent evidences show that they are pivotal part of gene regulatory networks including ceRNA networks (Jayarathna *et al*., 2021; P. Li *et al*., 2022; Z. Li *et al*., 2022), ultimately impacting cancer pathogenesis. As aberrant expression of pseudogenes has been reported in various cancer types (Emadi-Baygi *et al*., 2017; Hu, Yang and Mo, 2018), they may serve as potential diagnostic or prognostic markers.

A number of pseudogenes are recognized as potential cancer prognostic and diagnostic biomarkers, examples being *INTS6P1* with diagnostic value in hepatocellular cancer, *OCT4* related pseudogenes with prognostic value in multiple cancers and *PTENP1* with diagnostic value in gastric cancer and prognostic power in clear renal cell carcinoma (Poliseno, Marranci and Pandolfi, 2015). There are also a portion of studies on pseudogene-miRNA interactions. It is demonstrated that in gastric cancer, *PTENP1* exerts a tumor suppressive role by sponging miR-106b and miR-93, and thus hindering the downregulation of *PTEN* (Zhang *et al*., 2017). *CYP4Z2P* is another example, that by sponging miR-211, miR-125a-3p, miR-197, miR-1226, and miR-204 causes an increase in *CYP4Z1* translation which results in stimulation of tumor angiogenesis in breast cancer (Zheng *et al*., 2015). But how pseudogenes function in esophageal cancer related ceRNA networks along with lncRNAs, miRNAs and mRNAs is mostly unbeknown.

In the current study, an ESCA-related ceRNA network was constructed by using the data obtained from The Cancer Genome Atlas (TCGA). First, differentially-expressed mRNAs (DEmRNAs), differentially-expressed lncRNAs (DElncRNAs), differentially-expressed pseudogenes (DEpseudogenes) and differentially-expressed miRNAs (DEmiRNAs) were identified. Then, DEmRNAs, DElncRNAs and DEpseudogenes that were among the targets of DEmiRNAs were selected to create a ceRNA network. A protein-protein interaction (PPI) network for the DEmRNAs in the ceRNA network was furthermore constructed and the hub genes were determined. Survival analysis was carried out on the RNAs with degree centrality more than two in the ceRNA network. By considering hazard ratios of survival-related RNAs, synaptotagmin 10 (*SYT10*) was suggested as a potential prognostic biomarker in ESCA. The workflow of the present study is represented in fig 1.

**Fig 1.**
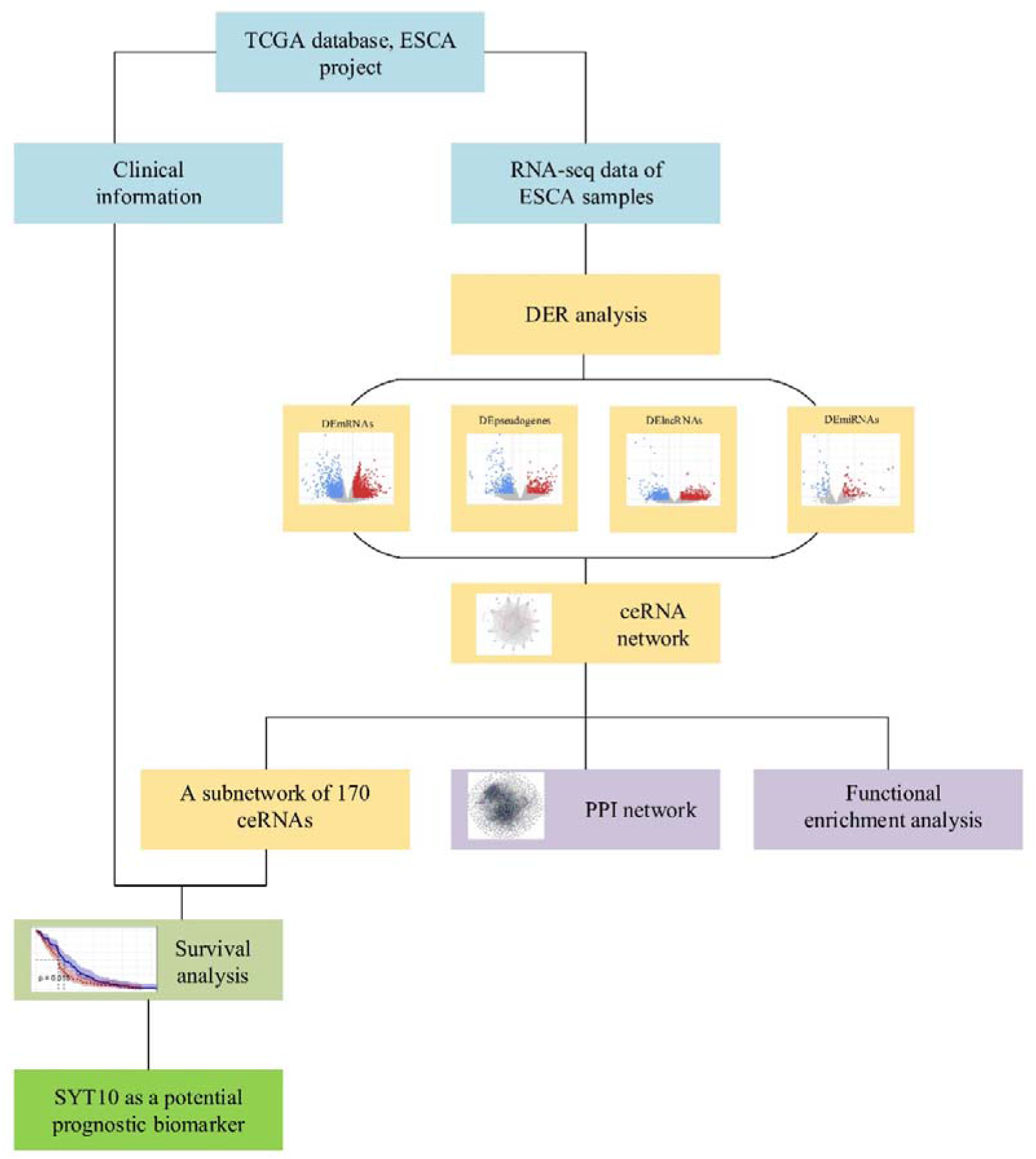
The workflow of the present study. ceRNA: competitive endogenous RNA, DER: differentially-expressed RNA, DEmRNAs: differentially-expressed mRNAs, DElncRNAs: differentially-expressed lncRNAs, DEpseudogenes: differentially-expressed pseudogenes, DEmiRNAs: differentially-expressed miRNAs, ESCA: esophageal carcinoma, PPI: protein-protein interaction network, SYT10: Synaptotagmin 10, TCGA: the cancer genome atlas

## Materials and methods

### Data retrieval and processing

RNA-seq data of 173 ESCA samples (162 tumoral and 11 normal) and corresponding clinical information along with miRNA-seq data of 199 samples (186 tumoral and 13 normal) was obtained from the TCGA database by using TCGAbiolinks package (Colaprico *et al*., 2015) in the RStudio software. The intersected data which contained 162 tumoral and 11 normal samples was normalized using the DESeq2 package (Love, Huber and Anders, 2014). DESeq2 package was then used for differential analysis. DERs were identified using |log_2_ fold change (FC)|>1 and false positive rate (FDR)<0.05 as thresholds.

### Construction of a ceRNA network

DEmRNAs, DEpseudogenes, DElncRNAs, and DEmiRNAs were used to construct a ceRNA network. Twenty percent of the most reliable miRNA-mRNA interactions was retrieved. To this aim, TargetScan, miRDB, and miRTarBase were selected and employed, using multiMiR package in R studio software (Hsu *et al*., 2011; McGeary *et al*., 2019; Chen and Wang, 2020; Huang *et al*., 2022). From TargetScan, interactions with a threshold of Context++ score≤-0.6, from miRTarBase, interactions with strong evidence and from miRDB, predicted targets with score more than 90% were acquired. Furthermore, the interactions between DEmiRNAs with DElncRNAs and DEpseudogenes were retrieved from RNAInter database with a score>0.5 (Kang *et al*., 2022). Finally, the ceRNA network was visualized in the Cytoscape software (version 3.9.3) (Shannon *et al*., 2003).

### PPI network construction

To inquire more into the DEmRNA interactions of the ceRNA network, a PPI network was established. The Search Tool for the Retrieval of Interacting Genes (STRING) database was used to construct a PPI network with minimum confidence score of 0.4 (Szklarczyk *et al*., 2019). Then, the aforementioned PPI network was exported to Cytoscape, where the genes with the highest degree centrality ranks were determined using cytoHubba plugin (Chin *et al*., 2014).

### Functional and Pathway Enrichment Analyses

To explore Gene Ontology (GO) and Kyoto Encyclopedia of Genes and Genomes (KEGG) pathway enriched terms in DEmRNAs of the ceRNA network, DAVID Bioinformatics Tool was employed (Huang, Sherman and Lempicki, 2009). The terms with FDR less than 0.05 were considered significant and illustrated in the R software using ggplot2 package.

### Survival analysis

From the ceRNA network, RNAs with degree centrality more than two were selected to create a subnetwork for further survival analysis. The association between the RNAs expression and patients’ overall survival was analyzed using survival and survminer packages in R. Samples were divided into two groups based on the median expression of each RNA, and Kaplan-Meier method was used to demonstrate the results. *P*. value<0.05 and hazard ratio≠1 was set as thresholds.

## Results

### Differentially-expressed RNAs in ESCA

The RNA-seq data obtained from TCGA database were analyzed and 3213 DEmRNAs (1433 down-regulated and 1780 up-regulated), 741 DEpseudogenes (443 down-regulated and 298 up-regulated), 1777 DElncRNAs (805 down-regulated and 972 up-regulated), and 173 DEmiRNAs (70 down-regulated and 103 up-regulated) were identified between 162 tumoral and 11 normal samples. The corresponding volcano plots are presented in fig 2.

**Fig 2.**
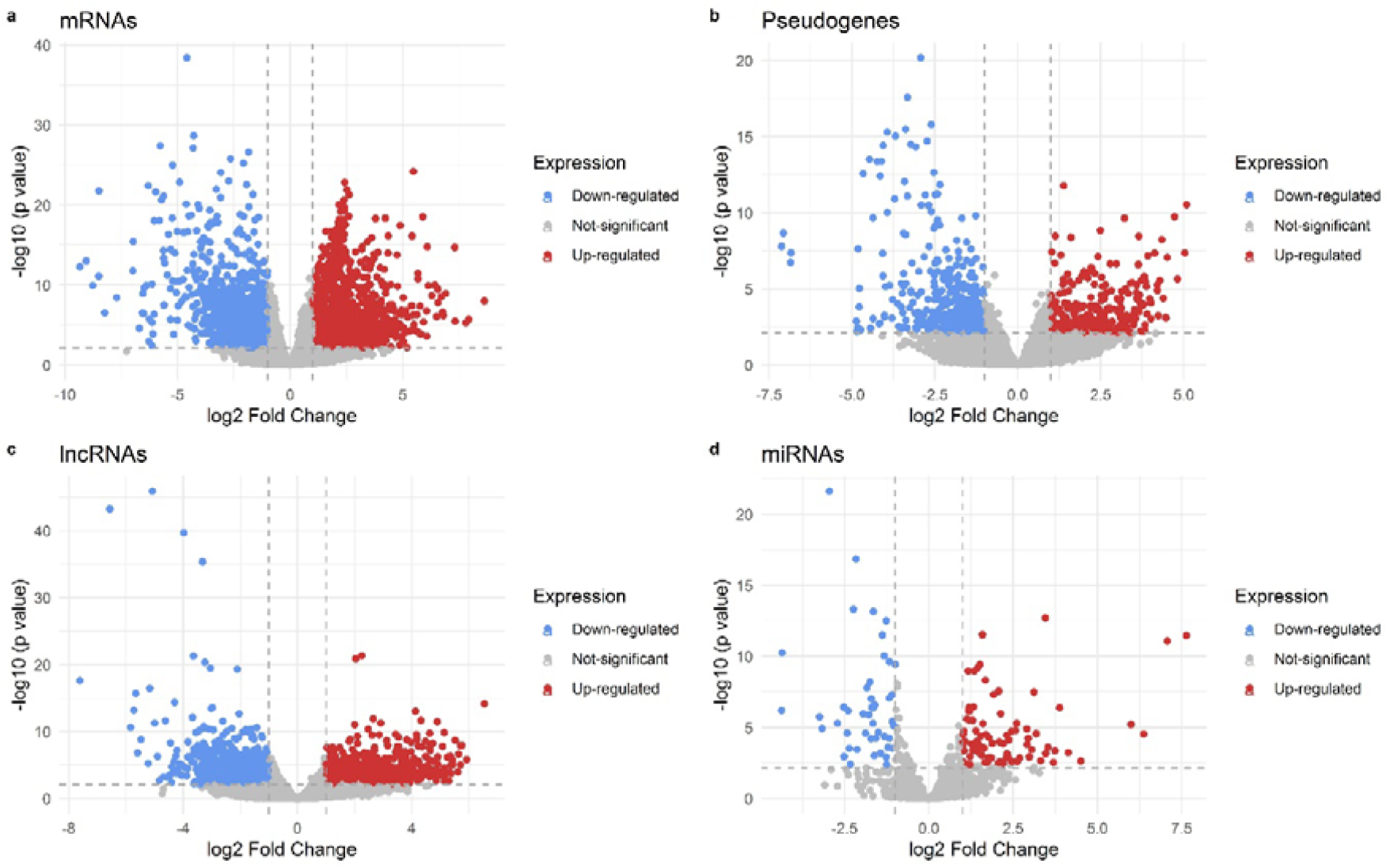
The volcano plots represent differentially-expressed RNAs between normal and tumoral tissues of ESCA. **a** Volcano plots of DEmRNAs; **b** DElncRNAs; **c** DEpseudogenes and **d** DEmiRNAs. |log_2_ FC|>1 and adjusted *p*-value<0.05 were considered as significant thresholds. Red dots stand for up-regulated RNAs, so do blue dots for down-regulated RNAs and gray dots show RNAs with not significant expression. ESCA: esophageal carcinoma, DEmRNA: differentially-expressed mRNAs, DElncRNAs: differentially-expressed lncRNAs, DEpseudogenes: differentially-expressed pseudogenes, DEmiRNAs: differentially-expressed miRNAs, log_2_ FC: log_2_ fold change

### ceRNA Network

To obtain the interactions between DEmRNAs and DEmiRNAs, three databases including TargetScan, miRTarBase, and miRDB were utilized by employing multiMiR package in the R software. There found to be 4337 total interactions between DEmRNAs and 195 unique DEmiRNAs. In the next step, the 195 unique DEmiRNAs were applied to RNAInter database to search for DElncRNAs-DEmiRNA and DEpseudogenes-DEmiRNAs interactions, and there found to be 80 and 40 interactions between DElncRNAs-DEmiRNA and DEpseudogenes-DEmiRNAs, respectively. Overall, with 730 discovered interactions, a ceRNA network was constructed which consisted of 502 DEmRNAs, 2 DEpseudogenes, 10 DElncRNAs and 15 miRNAs (Fig 3). An interactive version of the ceRNA network can be found on NDEx website (www.ndexbio.org/viewer/networks/0961f46b-f40d-11ed-aa50-005056ae23aa).

**Fig 3.**
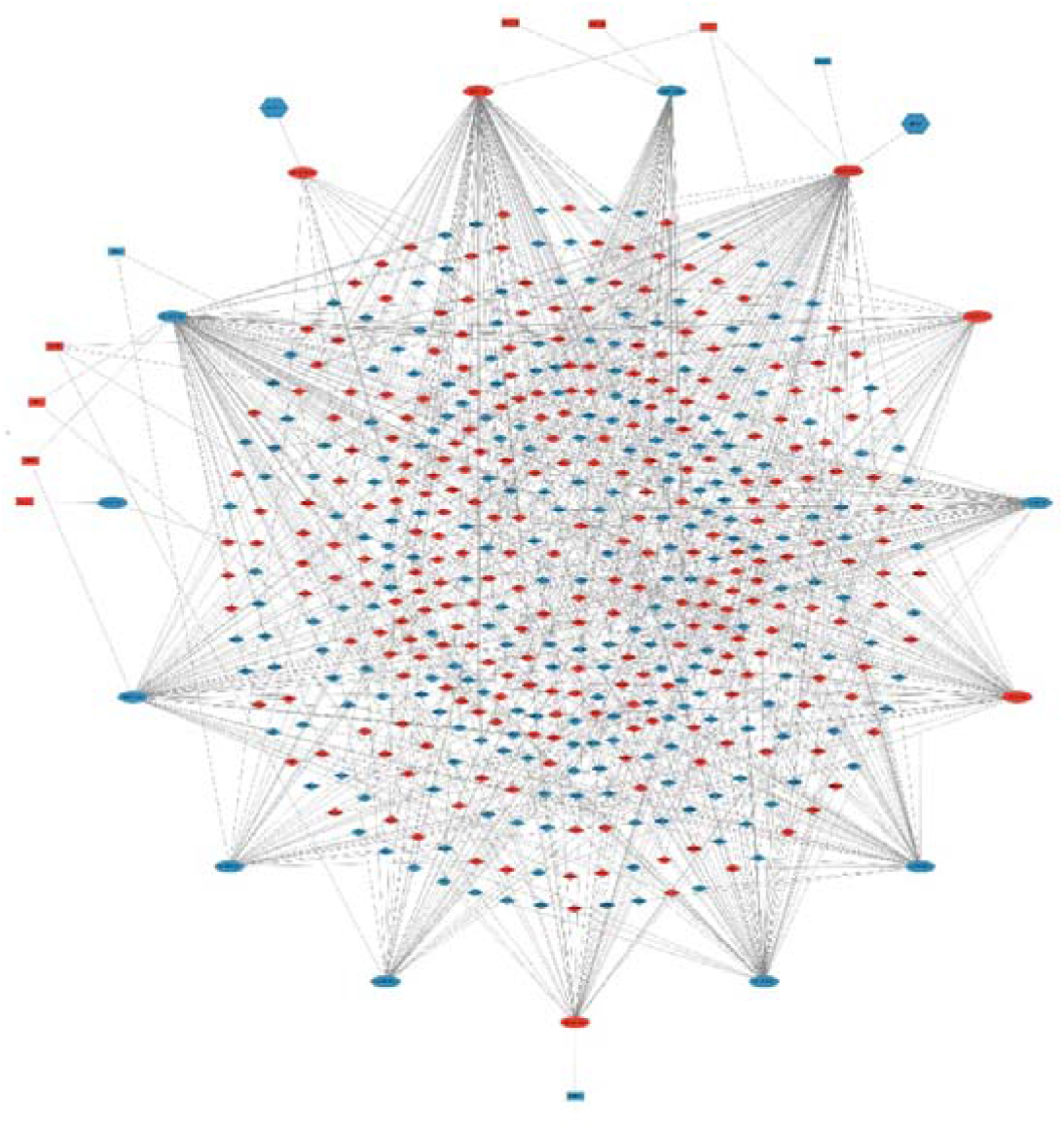
The ceRNA network of DEmRNA-DEpseudogene/DElncRNA-DEmiRNA in ESCA. Red and blue represent up-regulation and down-regulation, while diamonds, hexagons, rectangles and ellipses denote for DEmRNAs, DEpseudogenes, DElncRNAs and DEmiRNAs, respectively. ceRNA: competitive endogenous RNA, DElncRNAs: differentially-expressed lncRNAs, DEmRNA: differentially-expressed mRNAs, DEmiRNAs: differentially-expressed miRNAs, DEpseudogenes: differentially-expressed pseudogenes, ESCA: esophageal carcinoma

The network contained 529 nodes and 729 edges. Corresponding DEmiRNAs to 10 DElncRNAs and 2 DEpseudogenes are presented in tables 1 and 2, respectively. All DEmiRNAs and their matching DEmRNAs are presented in supplementary table 1.

**Table 1.** lncRNAs of the ceRNA network of ESCA and their associated miRNAs

| lncRNAs | Associated miRNAs |
| --- | --- |
| *AL158206.1* | hsa-miR-106b-5p |
| *CDKN2B-AS1* | hsa-miR-125a-5p |
| *FENDRR* | hsa-miR-18a-5p |
| *H19* | hsa-miR-106b-5p, hsa-miR-148a-3p |
| *HOTAIR* | hsa-miR-1-3p, hsa-miR-143-3p, hsa-miR-148a-3p, hsa-miR-7-5p |
| *HOXA11-AS* | hsa-miR-125a-5p |
| *LUCAT1* | hsa-miR-375-3p |
| *MEG3* | hsa-miR-21-5p, hsa-miR-664a-3p |
| *PVT1* | hsa-miR-195-5p, hsa-miR-26b-5p |
| *SNHG1* | hsa-miR-145-5p, hsa-miR-195-5p |

ceRNA: competitive endogenous RNA, ESCA: esophageal carcinoma, lncRNA: long non-coding RNA

**Table 2.**
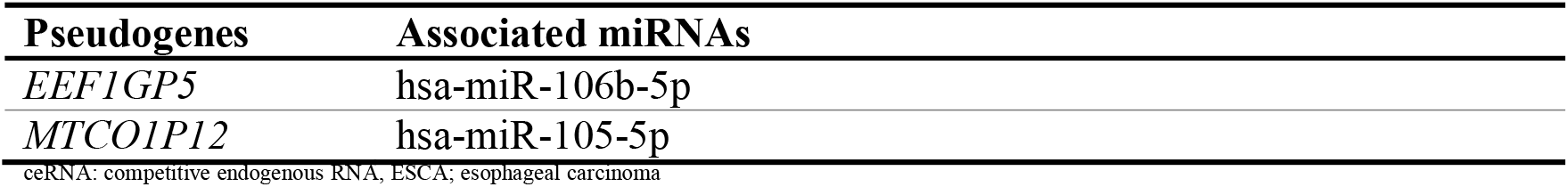
Pseudogenes of the ceRNA network of ESCA and their associated miRNAs

### PPI Network Construction and Functional Enrichment Analysis

To further investigate the interactions between the DEmRNAs of the ceRNA network, a PPI network was constructed with 501 nodes and 2518 edges (Fig 4a), in which, TNF, CCND1, FGF2, IL1B, MMP9, IGF1, TGFB1, CDK1, MMP2 and CXCL8 were identified as having the highest degree centralities (Fig 4b).

**Fig 4.**
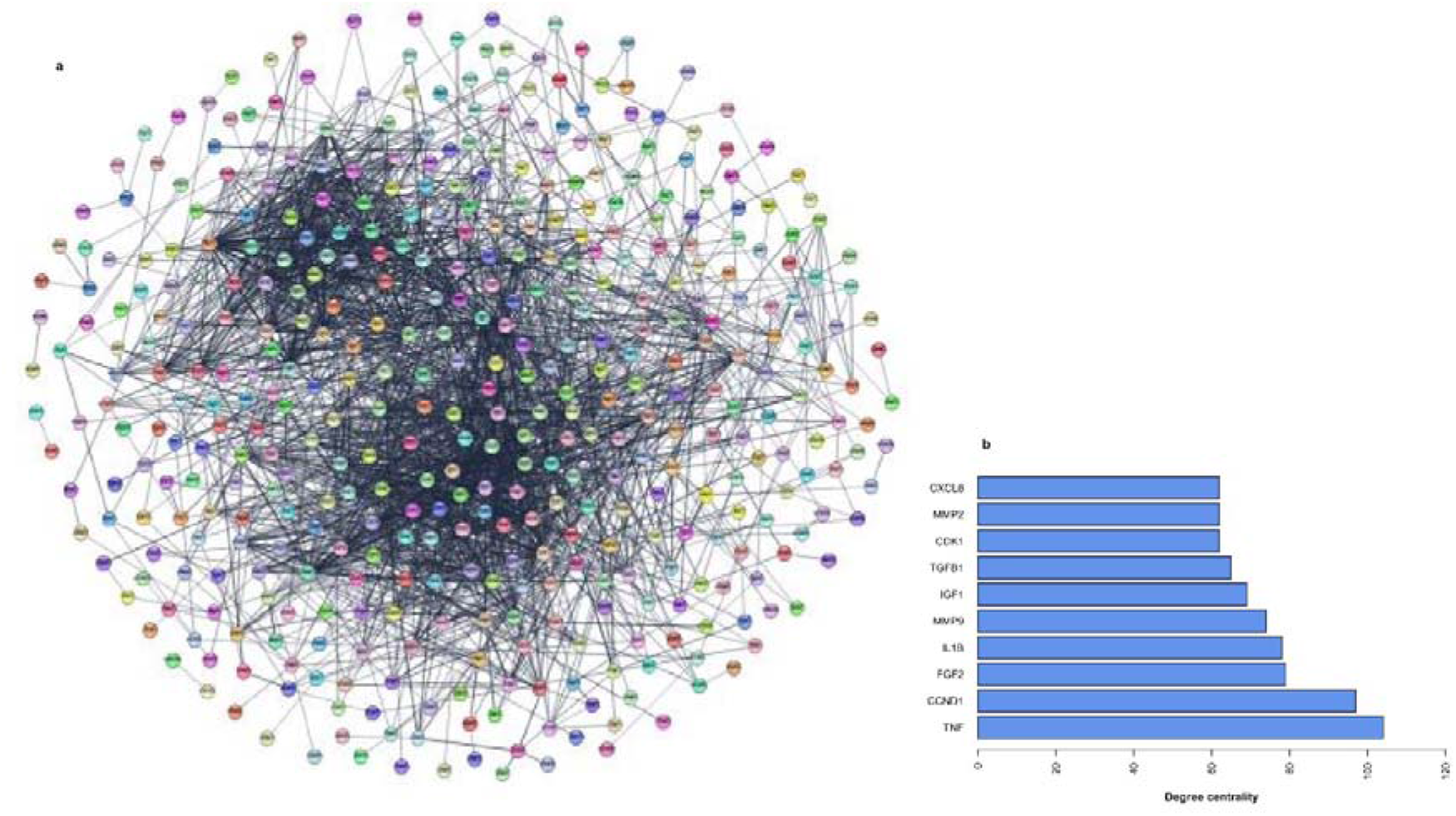
The PPI network of the DEmRNAs of the ceRNA network. **a** A PPI network was constructed with 501 nodes and 2518 edges. Nodes with no edges are not represented in the network; **b** Bar plot of the top ten nodes is ordered by network degree centrality. ceRNA: competitive endogenous RNA, DEmRNAs: differentially-expressed RNAs, PPI: protein-protein interaction

In the next step, to inquire into the functional analysis and cellular pathways in which the aforementioned DEmRNAs were enriched, GO and KEGG enrichment analyses were carried out. As the results show in fig 4a, most significant GO biological process terms include signal transduction, cell adhesion and positive regulation of gene expression. GO enrichment analysis also revealed that the associated proteins of DEmRNAs are mostly localized in plasma membrane and extracellular region/space (Fig 5a) and their related functions are that of sequence specific double strand DNA and receptor binding (Fig 5a). KEGG functional analysis also showed that ceRNA network–related DEmRNAs are mostly associated with pathways including neuroactive ligand-receptor interaction, cytokine-cytokine receptor interaction, Cell cycle, and protein digestion and absorption (Fig 5b).

**Fig 5.**
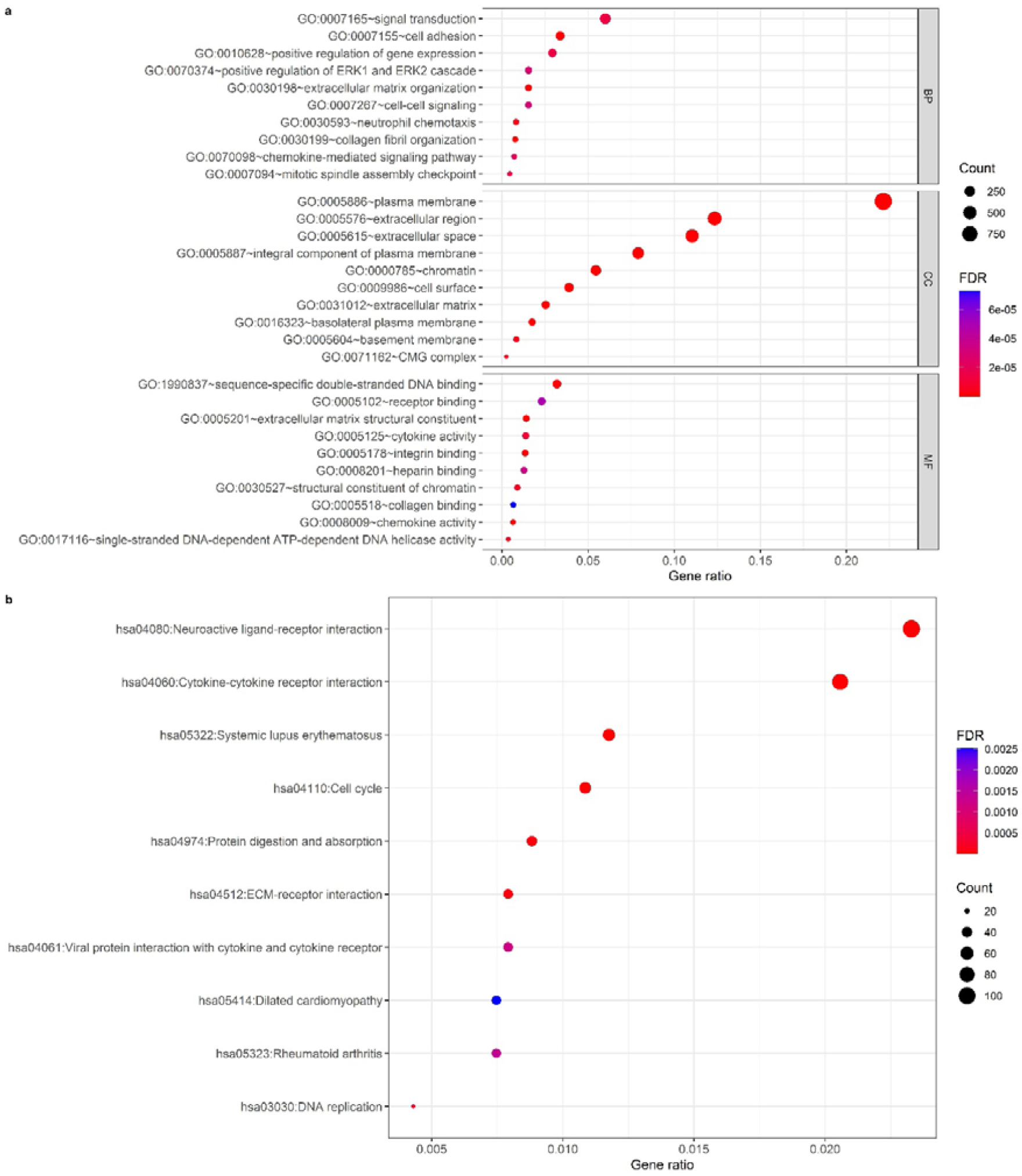
Dot plots of GO and KEGG functional enrichment analysis based on the DEmRNAs of the ceRNA network. **a** GO enrichment terms; **b** KEGG enrichment terms. The horizontal and vertical axes represent gene ratio and enrichment terms, respectively. For each term, FDR<0.05 was considered significant. DEmRNAs: differentially-expressed mRNAs, GO: Gene Ontology, FDR: false discovery rate, KEGG: Kyoto Encyclopedia of Genes and Genomes

### Survival Analysis of DERNAs in a ceRNA Subnetwork

A subnetwork of ceRNAs with each node having a degree centrality of more than two was selected to explore the association of ceRNAs with the patients’ overall survival. This subnetwork included 170 ceRNAs, and the ones significantly associated with overall survival were one DEmiRNA entitled hsa-miR-26b, and nine DEmRNAs including *SYT10, PVT1, DNMT1, COL4A1, ARHGEF28, MASTL, LRIG1, SULF1 and CDC25A*. Among these RNAs, those which their up-regulation was associated with poor overall survival included hsa-miR-26b, PVT1, DNMT1, COL4A1 and ARHGEF28, while those which their down-regulation was associated with poor overall survival were *SYT10, LRIG1, SULF1* and *CDC25A*. Among survival-related DEmRNAs, *SYT10, PVT1, DNMT1, COL4A1, ARHGEF28* and *MASTL* showed the highest hazard ratios (Table 3).

**Table 3.**
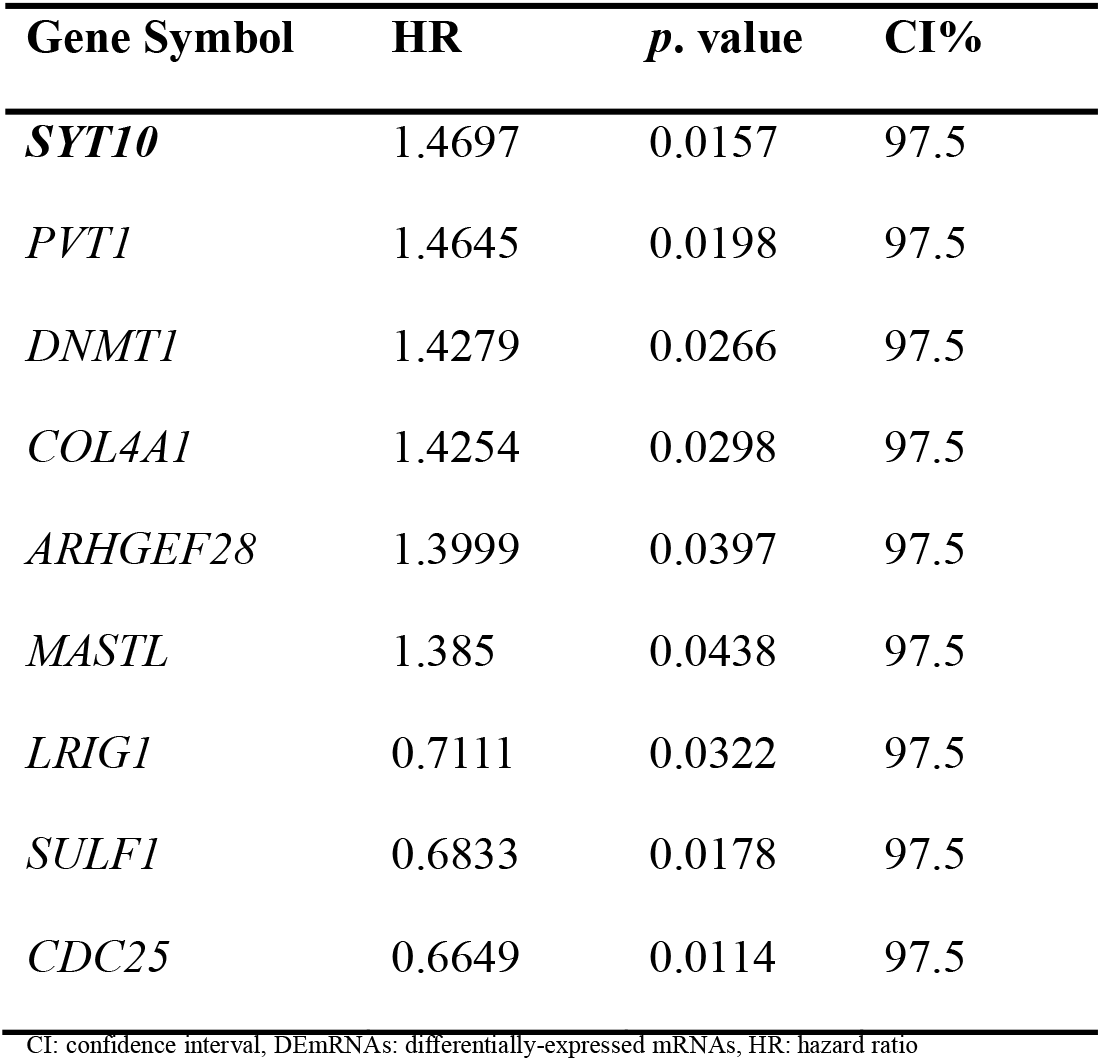
Survival-related DEmRNAs

Thus, it was assumed that these genes were of prognostic value. Kaplan-Meier curves of overall survival-related RNAs with the highest hazard ratios is presented in fig 6. Of note, none of the DElncRNAs in the subnetwork were associated with patients’ overall survival.

**Fig 6.**
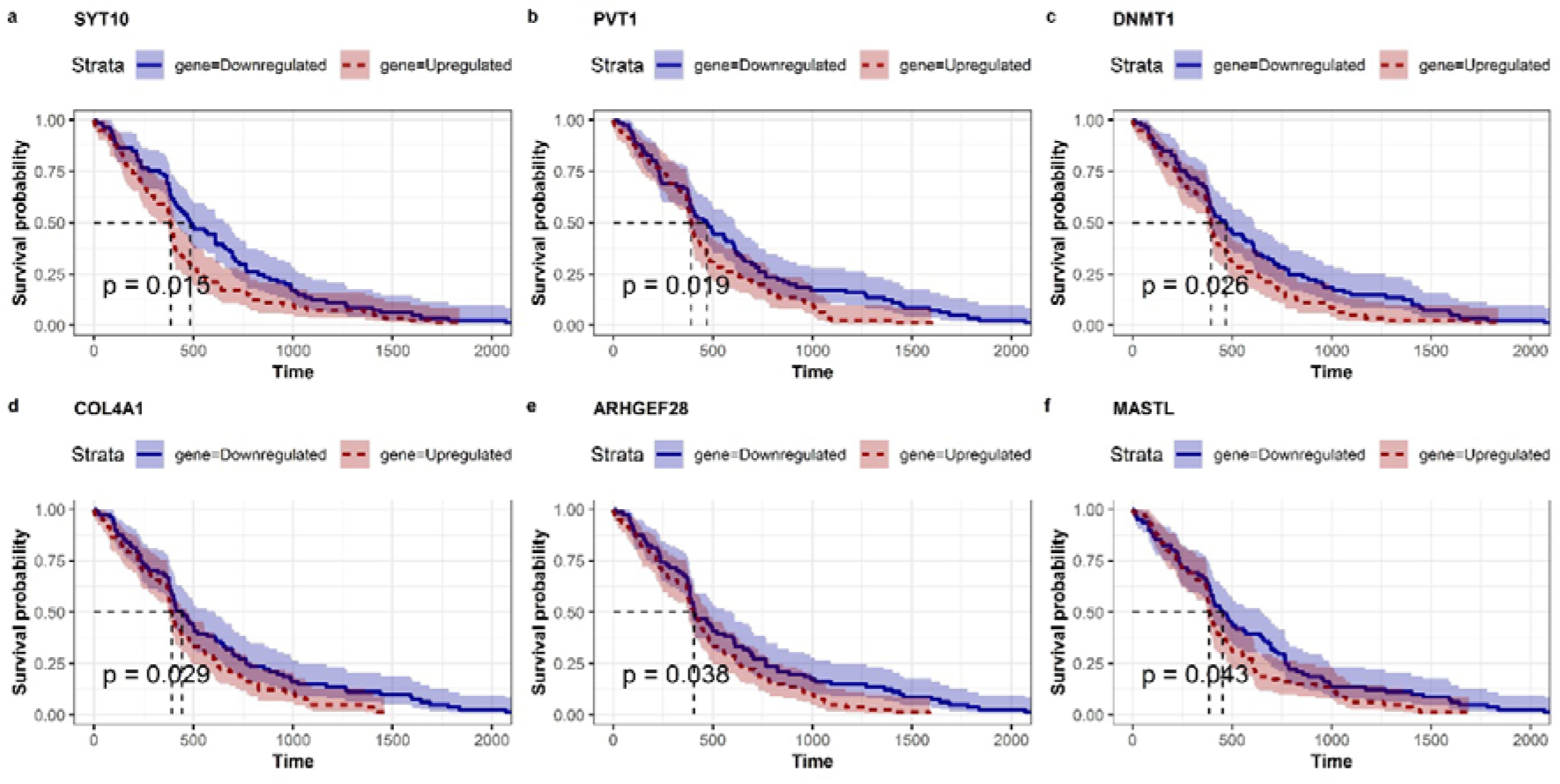
Kaplan–Meier survival curves of six overall survival-related RNAs with the highest hazard ratios. **a** Survival plot of *SYT10*; **b** *PVT1*; **c** *DNMT1*; **d** *COL4A1*; **e** *ARHGEF28*; **f** *MASTL* were constructed by division of ESCA tumor samples into two groups based on the median of gene expression. Red and blue colors stand for those samples with gene expression higher and lower than the median, respectively. ESCA: esophageal carcinoma

## Discussion

ESCA is one of deadliest cancers world-wide due to its aggressive nature and poor overall survival. Understanding the underlying ESCA tumorigenic molecular mechanisms can lead to identifying new cancer prognostic and diagnostic biomarkers, which can be used to boost patients’ quality of life. It has been clarified that long non-coding RNAs exert regulatory functions in cells and take part in tumorigenesis, but the details are yet to be shed light on (Schmitt and Chang, 2016). Amongst various classes of long non-coding RNAs, pseudogenes have gained a lot of attention, especially in the context of ceRNA regulatory networks, in which pseudogenes can talk to other RNAs harboring the same MREs, while miRNAs act as mediators (Hu, Yang and Mo, 2018).

To delve more in-depth into the characteristics of a ceRNA network in which pseudogenes are taken into account as one of the crucial components, this study was planned. To this aim, differentially-expressed mRNAs, pseudogenes, lncRNAs and miRNAs between ESCA tumoral and normal samples were identified. Based on predicted and validated interactions between 502 DEmRNAs, two DEpseudogenes, 10 DElncRNAs and 15 DEmiRNAs a ceRNA network was constructed which contained 529 nodes and 729 edges. To explore the interactions between DEmRNAs, a PPI network was constructed in which *TNF, CCND1, FGF2, IL1B, MMP9, IGF1, TGFB1, CDK1* and *MMP2* had the highest degree centralities. By analyzing GO and KEGG terms, it was deciphered that ceRNA network related DEmRNAs are active in signal transduction and cell adhesion, are mostly concentrated in plasma and extracellular region, and their significant GO molecular function terms include receptor and DNA binding. KEGG enriched terms also included neuroactive ligand-receptor interaction, cytokine-cytokine receptor interaction, cell cycle, neuroactive ligand-receptor interaction, and ECM-receptor interaction protein digestion and absorption.

Based on degree centrality ranks, a subset of ceRNA network was selected for survival analysis. The results showed that one DEmiRNA, which was hsa-mir-26b and nine DEmRNAs including *SYT10, PVT1, DNMT1, COL4A1, ARHGEF28, MASTL, LRIG1, SULF1* and *CDC25A* were associated with patients’ overall survival. By considering the hazard ratios, it was concluded that *SYT10* may be a potential prognostic biomarker for ESCA.

In human, synaptotagmin family is constituted of 17 isoforms (SYT1 to SYT17) which mostly participate in calcium-dependent membrane fusion regulation (Wolfes and Dean, 2020). There are a number of ways by which cell communication is conducted, and the most recent one that has gained attention is by extracellular vesicles that act as means of signaling (van Niel, D’Angelo and Raposo, 2018). Exosomes are secreted by almost all types of cells including tumoral ones, and can be found in many body fluids. Signals transmitted by tumor-derived exosomes result in manifestation of many of the cancer characteristics, including metastasis and drug-resistance (Tai *et al*., 2018). Synaptotagmin family members are found to take part in exosome release, for example it was shown that knockdown of *SYT7* can result in the decreased release of exosomes along with the decrease in numbers and activity of invadopodia in head and neck squamous cell carcinoma (HNSCC) cell lines (Hoshino *et al*., 2013). In 2017, *SYT7* was reported for the first time as an oncogene in colorectal cancer (Wang *et al*., 2018), a notion which was supported afterwards by many studies, suggesting that *SYT7* contributes to cellular pathway regulation in many cancer types resulting in cancer progression, for example in lung cancer, melanoma, HNSCC and thyroid cancer (Fei *et al*., 2019; Chu, Wan and Zhang, 2021; Fu *et al*., 2021; Dong *et al*., 2022).

*SYT13* is another member of synaptotagmin family which doesn’t share the Ca^2+^ binding site with other prominent members, including *SYT1, SYT2* and *SYT3*, but still participates in vesicle trafficking. Lack of function of *SYT13* contributes to impaired endocrine cell egression, whereas its’ gain of function results in reduced cell-matrix adhesion *in vitro* (Bakhti *et al*., 2022). Over expression of *SYT13* is also associated with cancer advancement; it is shown that patients with gastric cancer metastasis or peritoneal recurrence have higher levels of *SYT13*, and knockdown of *SYT13* in gastric cancer cell lines results in reduced cell migration and invasion (Kanda *et al*., 2018).

There are evidences of syanptotagmin family members’ contribution to poor prognosis in ESCA. For example, in 2022, Lian *at al* introduced a hypoxia- and immune-associated prognosis signature for ECSA which contained eight RNAs, one of them being *SYT1* (Lian *et al*., 2022). Another proof of the significance of the syanptotagmin family members in ESCA progression and poor survival, is the effect of hypoxia as a hallmark of cancer (Ruan, Song and Ouyang, 2009) has in both development of malignancies and exosome secretion. As He *et al* suggest in (He *et al*., 2022), hypoxia can cause overexpression of synaptotagmin family members and other related factors, resulting in boosted exosome release, which leads to cancer progression. There are also accumulating studies suggesting that hypoxia may be a crucial factor with a role in cancer drug resistance in cancers like ESCA (Zhu *et al*., 2021).

*SYT10* contributes to secretion of insulin-like growth factor 1 in olfactory bulb neurons. Furthermore, Syt10 is known to be up-regulated by neuronal activity which protects neurons against excitotoxicity induced by pathophysiological synaptic activity (Wolfes and Dean, 2020). *SYT10* has been introduced as a candidate gene responsible for cisplatin resistance in two esophageal squamous cell carcinomas cell lines (Tsutsui *et al*., 2015). In the ceRNA network that was constructed in the present study, *SYT10* was related to two miRNAs, including hsa-mir-195-5p and hsa-mir-26b-5p, both of which act against epithelial mesenchymal transition (EMT) and proliferation of the cancer cells (Ma *et al*., 2021; Zhou *et al*., 2021).

There is not much evidence of *SYT10* taking any major part in cancer proliferation and cellular pathway regulation leading to tumorigenesis, but as previous studies suggest (Tsutsui *et al*., 2015; Zhu *et al*., 2021; He *et al*., 2022), *SYT10*, as a syanptotagmin family member, may play a role in drug resistance in ESCA and thus contribute to poor survival and prognosis. A detailed recent review study on potential roles of SYT family members in cancers has been properly described elsewhere (Suo, Xiao and Wang, 2022).

## Conclusion

In conclusion, we constructed a ceRNA network by analyzing the mRNA, pseudogene, lncRNA and miRNA expression profiles of esophageal cancer from the TCGA cohort. We identified 6 mRNAs which their higher expression was associated with poor overall survival. Among these, *SYT10* had the highest hazard ratio which can be considered as a potential prognostic biomarker in ESCA. Further experimental evaluations and clinical validations are needed for approval of this candidate biomarker.

## Supporting information

Suppl. Table 1. Differentially-expressed miRNAs and their matching differentially-expressed mRNAs

## Data Availability

The results published or shown here are in whole or part based upon data generated by the TCGA Research Network (https://portal.gdc.cancer.gov/legacy-archive/search/f). Data are available upon request to the corresponding author.

## Statements & Declarations

### Funding

This work was partially supported by a grant from Isfahan University of Medical Sciences, Isfahan, Iran [grant number 3400942].

### Competing Interests

The authors declare that there are no relevant financial or non-financial interests to disclose.

### Author Contributions

Milad Daneshmand-Parsa: Data Curation (equal); Formal Analysis (equal); Investigation (equal); Methodology (equal); Software (equal); Writing – Original Draft Preparation (equal); Writing – Review & Editing (equal)

Sharareh Mahmoudian-Hamedani: Data Curation (equal); Formal Analysis (equal); Investigation (equal); Methodology (equal); Software (equal); Visualization (lead); Writing – Original Draft Preparation (equal); Writing – Review & Editing (equal)

Parvaneh Nikpour: Conceptualization (lead); Funding Acquisition (lead); Investigation (equal); Project Administration (lead); Resources (lead); Supervision (lead); Validation (lead); Writing – Original Draft Preparation (equal); Writing – Review & Editing (equal)

## Notes

### Competing Interest Statement

The authors have declared no competing interest.

